# Systematic evaluation of incomplete penetrance in 4,325 rare disease families reveals divergent transcriptional networks in *CHD8* pathology

**DOI:** 10.64898/2026.10.05.26364218

**Authors:** Mugdha Singh, Sarah L. Stenton, Emily Groopman, Moriel Singer-Berk, Jialan Ma, Sanaa Choufani, Emily O’Heir, Stephanie DiTroia, Ikeoluwa Osei-Owusu, Lynn Pais, Gabrielle Lemire, Melanie O’Leary, Christina Austin-Tse, Casie A. Genetti, Katherine N. Anderson, Monica H. Wojcik, Alan H. Beggs, Jennifer E. Neil, Abbe Lai, Christopher A. Walsh, Julie A. Jurgens, Brenda J. Barry, Elizabeth C. Engle, Sandra Donkervoort, Carsten G. Bönnemann, Emanuela Argilli, Elliott H. Sherr, Patricia Kipkemoi, Zandre Bruwer, Kirsten A. Donald, Volker Straub, Ana Topf, Susan M. White, Tiong Yang Tan, Lyndon Gallacher, Peers Arts, Matilda R. Jackson, Thuong T. Ha, Christopher P. Barnett, Alicia B. Byrne, Hamish S. Scott, Vijay S. Ganesh, Heidi L. Rehm, Rosanna Weksberg, Anne O’Donnell-Luria, Sanna Gudmundsson

**Author notes:** corresponding author **Corresponding authors:** Sanna Gudmundsson, Anne O’Donnell-Luria. shared authorship (Anne O’Donnell-Luria, Sanna Gudmundsson).

## Abstract

**INTRODUCTION:** Despite major advances in genomic sequencing and analytical pipelines for rare disorders, more than 50% of affected individuals remain without a molecular diagnosis. One challenge is the interpretation of inherited variants exhibiting incomplete penetrance (IP).

**MATERIALS AND METHODS:** We developed a pipeline to systematically investigate inherited variants as the cause of rare disease in 4,325 families, consisting of affected probands and unaffected parents, from the Broad Center for Mendelian Genomics, part of the GREGoR Consortium. As a proof-of-concept of further omics follow-up, epigenomic and transcriptomic analyses were performed in one family harboring an inherited disease-causing frameshift variant in *CHD8*.

**RESULTS:** We identified 81 families where the affected proband harbored a putative disease-causing variant inherited from a reportedly unaffected parent. Clinical re-evaluation identified 30 families in which an inherited variant was considered diagnostic, including six previously missed diagnoses. In this cohort, inherited IP variants account for 1.6% of the diagnosed cases, with an overall diagnostic yield of 0.4% in the cohort. Methylation analysis of the *CHD8* family confirmed a positive *CHD8* clinical episignature in the male proband, but also in the reportedly unaffected mother. Transcriptomic analysis showed that despite similar *CHD8* haploinsufficiency, the proband and mother displayed divergent downstream transcriptional effects.

**DISCUSSION:** This study highlights the importance of considering parentally inherited IP variants in rare disease, with at least 1 in 60 diagnostic variants inherited from a reportedly unaffected parent, and demonstrates the potential of integrating omics evaluation to improve variant interpretation.

## Introduction

Despite major advances in genomic sequencing and analytical pipelines for rare disorders, more than 50% of affected individuals remain without a molecular diagnosis.^1–3^ The current diagnostic gap reflects challenges in interpreting variant effects and their association with disease.^4–6^ One major challenge is interpreting inherited variants that exhibit incomplete penetrance (IP; absence of phenotype) or variable expressivity (VE; variability in features or severity of phenotype). Highly discordant phenotypes among family members sharing the same variant complicate diagnosis and genetic counseling, both by raising questions regarding the pathogenicity of the disease-causing inherited variant and about future risk of clinical manifestations in currently unaffected variant-carrying family members.^7,8^

The emergence of large-scale cohorts has enabled systematic investigation of IP. Population studies have improved understanding of penetrance prevalence by providing estimates for disorders such as prion disease,^9^ Turner syndrome,^10^ maturity-onset diabetes of young (MODY),^11,12^ osteogenesis imperfecta,^13,14^ and inherited retinal diseases.^15,16^ Beyond penetrance estimates, genomic analyses of a broader range of rare disease variants in population data, as well as statistical variant-phenotype association studies using, for example, the UK Biobank, confirm that IP/VE poses a challenge in clinical diagnostics.^17–20^ Several factors have been suggested to drive IP/VE, including polygenic background,^21–24^ modifier variants,^25–27^ somatic variants,^28^ monoallelic expression,^29^ transcript level modifiers,^30,31^ sex-specific factors^32^ and digenic inheritance.^33^ Our previous population-scale work on IP across 77 severe haploinsufficiency disorders in 76,215 genomes from the Genome Aggregation Database (gnomAD) reported an explanation for the absence of disease in 95% of pLoF variants investigated. In line with previous reports, alternative splicing was suggested to contribute to the reduced effect of pLoF and a potential driver of incomplete penetrance. However, 5% of pLoF variants could not be explained by genomic investigation alone, underlining the limitations of genomic investigation in understanding IP.^19^

Recent studies have dived deeper into the molecular mechanisms of IP, including mapping of common alleles in trans with a variant causing *FOXI3*-associated craniofacial microsomia,^34^ common regulatory variation in *TYR* causing albinism,^35^ as well as an epistatic interaction between a rare *SMAD6* variant and common *BMP2* variant in craniosynostosis^36^. Although few and disease-specific, these examples highlight the value of molecular follow-up for understanding drivers of IP. Omics data, including transcriptomic and epigenomic profiles, can provide direct evidence to explain inter-individual differences in variant effects, improving genome interpretation and offering a powerful approach for investigating IP. However, because large rare disease cohorts often have limited phenotypic information and few opportunities for longitudinal re-phenotyping, broader assessments of the contribution of inherited IP to rare disease have been lacking.

In this study, we map variants inherited from reportedly unaffected parents as the cause of rare disease in their affected offspring in 4,325 families enrolled in the Broad Center for Mendelian Genomics (CMG) project **(Figure 1)**. Following detailed clinical review, 30 inherited variants in 30 families were determined to be diagnostic, including six previously missed diagnoses. Further omics investigation of a family with a pathogenic *CHD8* variant and intrafamilial phenotypic variability, with severe symptoms in the proband and mild clinical features in the variant-carrying mother, revealed a positive episignature for both but distinct transcriptomic profiles.

**Figure 1:**
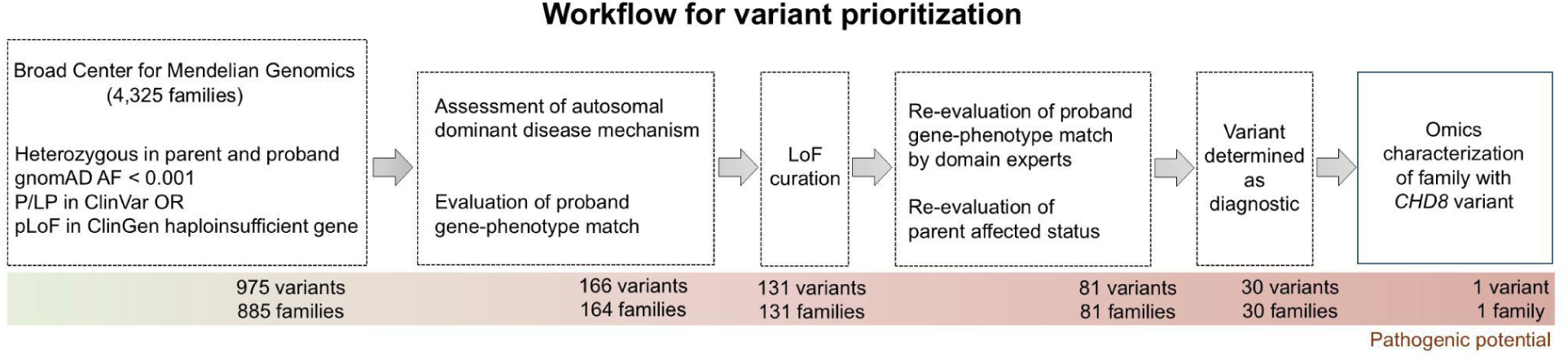
Overview of the variant prioritization pipeline. Applied filters, literature review, LoF curation and clinical assessments reduced 975 variants from 885 families to 30 diagnostic variants in 30 families. A family with a *CHD8* variant was selected for additional transcriptomic and epigenomic characterization. Variant and family counts at each stage are shown beneath the workflow.

## Methods

### Ethical approval

All participants provided informed consent before participating in this study. This study was approved by the Mass General Brigham Institutional Review Board (protocols 2016P001422 and 2013P001477).

### Sequencing of rare disease families

The study includes 4,325 families from the Broad Institute Center for Mendelian Genomics (CMG) cohort, many of which are part of the Genomics Research to Elucidate the Genetics of Rare Disease (GREGoR) consortium dataset, that have at least one reportedly unaffected enrolled parent. Genome sequencing (GS; n = 946) and exome sequencing (ES; n = 3,379) were performed using Illumina platforms (Illumina Inc, San Diego, CA) to generate 150-bp paired-end reads. GS was generated from PCR-free libraries sequenced with HiSeq 2500 or HiSeq X Ten v2 (Illumina Inc) with mean coverage >30×. ES employed Nextera (Illumina Inc) or Twist capture (Twist Bioscience, South Francisco, ∼38 Mb), achieving >80% at ≥20× and a mean coverage of >60×. Reads were aligned to GRCh38 with BWA, followed by base quality score recalibration and SNV/indel calling using GATK best practices (v4.0; Broad Institute, Cambridge, MA). Genome sequence data was reprocessed with DRAGEN v3.7.8 before depositing it to dbGaP/AnVIL.

### Variant filtering

Variants were filtered to retain those with allelic balance ≥ 0.2, genotype quality ≥ 40, and sequencing depth ≥ 10 reads. Among these, we included inherited heterozygous ultra-rare (gnomAD v2.1 or v3.1 allele frequency ≤ 0.001; available version at the time of analysis) variants, classified as pathogenic or likely pathogenic (P/LP) in ClinVar or annotated as predicted loss-of-function (pLoF) in a gene reported as haploinsufficient by ClinGen’s gene dosage sensitivity curation (accessed on November 13^th^, 2022).^37^ Each case was analyzed using the *seqr* platform (Broad Institute)^38^ and manual literature review focusing on the variants’ consistency with autosomal dominant disease mechanisms, as well as concordance between proband clinical features and known disease phenotypes reported in the literature and OMIM.^39^ pLoF variants were curated as described previously.^40^ Rules for LoF curation are specified in **Supplemental Table 1(a-b)**. SpliceAI was employed to predict splicing consequences^41^ using scores calculated with SpliceAI Lookup employing a cutoff of ≥ 0.2 (https://spliceailookup.broadinstitute.org/). The refined set of candidate variants in 81 families were evaluated by the individual’s primary clinical research team, including 24 clinical and domain-expert groups, to reassess clinical fit in the proband and assess the phenotype of the reportedly unaffected variant-carrying parent.

### *CHD8* methylation analysis

The episignature for the *CHD8* (HGNC:20513) family (RGP_526; proband and mother) was generated using the Illumina MethylationEPIC v2 array (Illumina Inc; 930,207 probes) from peripheral blood-derived DNA. Support vector machine (svm) classification scores were calculated for the proband and mother as described previously.^42^ For independent validation, 94 of 103 validated *CHD8* episignature probes overlapping between the family data and publicly available blood methylation reference data (GEO: GSE113967; Illumina Methylation 450 BeadChip; Illumina Inc; n = 7 *CHD8* cases, n = 48 controls) were used for classification and visualization as previously described.^42^ Briefly, median DNA methylation was computed at each of the 94 episignature probes across cases and controls to generate control and reference *CHD8* profiles. A *CHD8* classification score was defined as:

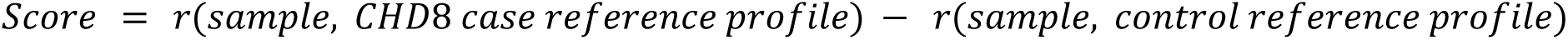

where r is the Pearson correlation coefficient.^43^ An individual with a positive score was classified as positive for the *CHD8* episignature, whereas an individual with a negative score was classified as negative. For the PCA analysis, beta values (representing DNA methylation at a CpG probe) across the 94 *CHD8* signature probes were normalized before performing principal component analysis (PCA) using scikit-learn Python module.^44^ For each probe, the normalized value was obtained by subtracting the beta with the mean beta value across all the samples, divided by the standard deviation.

### RNA sequencing analysis

For the family (RGP_526) with the *CHD8* frameshift variant, short-read RNA sequencing of lymphoblastoid cell lines (LCLs) from the mother and proband was performed at the Broad Institute. We used a polyA-tail RNA selection kit (Illumina TrueSeq Stranded mRNA; Illumina Inc.), generating 150 million 150 bp paired reads per sample, data was processed according to previously published protocols.^45^ Transcripts per million (TPM) read counts were generated using RNA-SeqQc v2.4.211 (Broad Institute) and the quality matrices were summarized with MultiQC. OUTRIDER^46^ analysis was performed for transcriptome-wide expression outlier analysis. Genome-wide distribution of expression outliers was assessed using a sliding window approach (5Mb window, 1Mb step) across all autosomes, counting significant outliers (p-adjust <0.05) per window per individual for proband, mother and controls (n = 105, including 100 GTEx and 5 internal RNA data from LCLs). The number of deduplicated outlier genes within the 6p21.3 locus (chr6:28,510,120-33,480,577) was compared between the *CHD8* mother, proband and controls using empirical p-values, calculated as the proportion of control individuals with outlier gene counts greater than or equal to the observed value. For the observations exceeding all controls, p-values are reported as p<1/(n+1) following Laplace correction.

### Gene set enrichment analysis (GSEA)

To test whether outlier signature reflected broader dysregulation of neurodevelopmental disorder (NDD) networks, we performed preranked gene set enrichment analysis (GSEA) using clusterProfiler R package. For both the proband and mother, the complete set of expressed genes (n = 12,419) was preranked by OUTRIDER Z-scores, calculated relative to 105 controls. These ranked lists were queried against curated autism associated genes that were obtained from the Simons Foundation Autism Research Initiative (SFARI) database (gene.sfari.org, 2026 release) and clinical phenotype specific from Human Phenotype Ontology (HPO) gene sets. Both Category 1 alone (n = 244, strongest with most replicated genetic evidence) and Combined Categories (1+2) (n = 710 strong candidates in category 2, for a combined n = 952; 3 genes without mappable Entrez IDs were excluded) were used as GSEA sets in this study.^47^ Clinical phenotype-specific gene sets were retrieved via the msigdbr R package from HPO, specifically targeting annotations for autism (n = 207), macrocephaly (n = 112), and intellectual disability (n = 738). The GSEA algorithm calculated a running enrichment score (ES) across ranked gene lists, with normalized enrichment score (NES) reflecting magnitude and direction of coordinated gene set dysregulation. Statistical significance was evaluated using a false discovery rate (FDR) threshold determined via 1,000 gene-label permutations (seed = 42).

### Polygenic risk estimation

Polygenic risk scores (PRS) for the mother and proband (RGP_526) carrying a pathogenic *CHD8* variant were calculated. PRS was derived using published SNP weights from the genome-wide ASD PRS (PGS000327)^48^, comprising 35,087 variants. PRS was computed using the weighted allele count method by summing the product of effect allele dosage and published effect sizes, without p-value thresholding or linkage disequilibrium pruning. The scoring file was harmonized to GRCh38 using pre-computed coordinates from the PGS catalog. Variants on non-autosomal contigs, duplicate loci and position lacking genomic coordinates were excluded, leaving 35,0074 autosomal variants. Effect allele orientation was verified against the family VCF prior to genotype extraction using the pysam. Genotypes were successfully obtained for 34,677 variants (98.9%). PRS were calculated using 34,165 variants shared between the family VCF and 1000 Genome Phase 3 reference panel. The scores were evaluated relative to ancestry-matched individuals from the 1000 Genome Project Phase 3 by subtracting the reference mean and dividing by the reference standard deviation.

## Results

### Investigation of 4,325 families yielded 131 inherited candidate incompletely penetrant variants

We developed a systematic variant prioritization workflow to identify candidate inherited variants that might be disease-causing in the probands but demonstrate IP in the reportedly unaffected parent. The workflow was applied to 4,325 parent-child pairs (families) from the Broad CMG cohort, including 946 families with genome sequencing and 3,379 families with exome sequencing. We filtered for inherited, heterozygous ultra-rare variants (allele frequency ≤ 0.001 in gnomAD) and either pathogenic or likely pathogenic (P/LP) in ClinVar or predicted loss of function (pLoF) in a gene reported as haploinsufficient by ClinGen.^37^ This filtering strategy identified 975 variants across 885 families **(Figure 2a)**. Of 975 initial candidate variants, 68.6% were pLoF (n = 669), 26.8% missense (n = 262), 2.9% non-coding (n = 29), 0.82% synonymous (n = 8, all predicted to impact splicing), and 0.72% in-frame indels (n = 7). Overall, 70% of the variants were P/LP in ClinVar, and the remaining 30% lacked ClinVar germline classification but were pLoF variants in haploinsufficient genes. Of 478 genes, 50.4% (n = 241) harbored a single variant, 24.7% (n = 118) two variants, and 24.9% (n = 119) had more than two variants (**Supplemental Figure 1)**. The 975 filtered variants were carefully evaluated for consistency with an autosomal dominant disease mechanism, as well as concordance between the proband’s phenotypes and previous literature reports of clinical manifestations, returning 166 variants in 164 families. The most common reason for exclusion was that the variant was reported to cause autosomal recessive disease in a gene associated with a condition with both dominant and recessive inheritance, and the parent and child were thus heterozygous carriers of the autosomal recessive trait **(Supplemental Figure 2)**.

**Figure 2:**
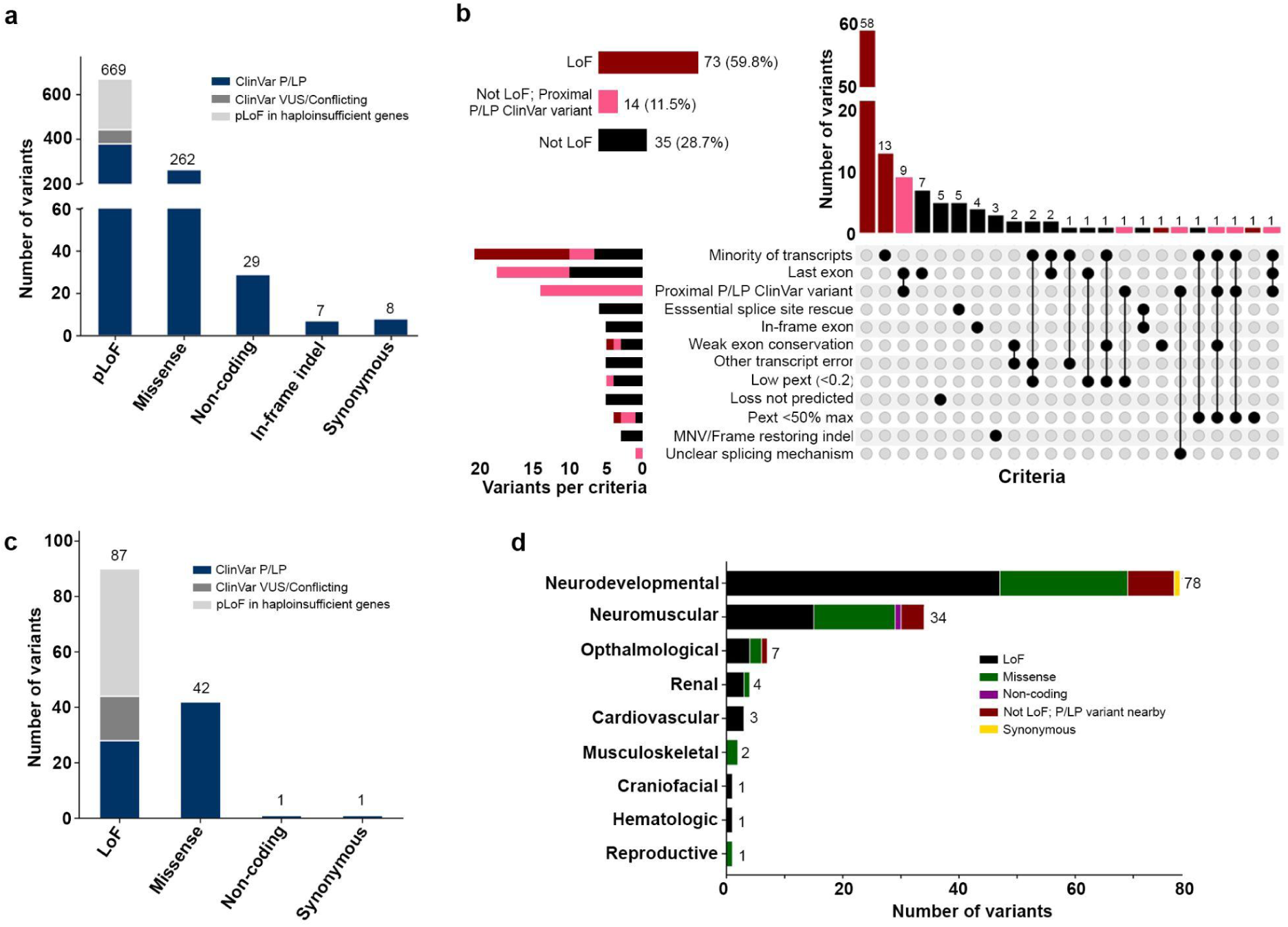
Analysis of 4,325 families from the Broad CMG cohort yielded 131 inherited candidate variants for incomplete penetrance (IP). **(a)** Barplot with 975 filtered variants, with the number of variants per variant type colored by P/LP: Pathogenic/likely pathogenic (blue); VUS: Variant of uncertain significance (dark grey); pLoF: predicted loss of function variant (light grey). **(b)** UpSet plot depicting pLoF curation results across 122 pLoF variants. Each row in the lower matrix represents the curation criteria resulting in a Not pLoF verdict. **(c)** Barplot with 131 candidate variants remaining after filtering for disease mechanism, phenotype overlap, and pLoF curation, colored as in 2a. **(d)** Horizontal barplot depicting the distribution of the 131 candidate IP variants across nine disease groups in the Broad CMG cohort, colored by variant type.

Of the 166 candidate variants, 122 were pLoF, and all 122 pLoF variants were further assessed to determine whether they were predicted to result in protein loss due to nonsense mediated decay (NMD)^40^ or escape NMD. Of 122 pLoF variants, 60% (n = 73) were predicted as NMD prone and classified as LoF, whereas 40% (n = 49) were predicted to escape NMD and classified as Not LoF. Of the 49 Not LoF variants, 28.5% (n = 14) had at least one reported ClinVar P/LP pLoF variant downstream or nearby indicating protein truncation may still result in disease. For the 35 Not LoF variants that were excluded, 20% (n = 7) were excluded based on being located in the last exon or 50 bp of the penultimate exon, 14% (n = 5) because of a predicted in-frame splice rescue within +/-1-2 codons, 14% (n = 5) because in the absence of functional data the impact on splicing was not supported by SpliceAI, 11% (n = 4) because SpliceAI predicted splicing of an in-frame exon, 9% (n = 3) because presence of a multi-nucleotide variant (MNV) or frame-restoring indel, and the remaining 32% (n = 11) due to a combination of listed criteria including a low proportion expressed across transcripts (pext) score of < 0.2 **(Figure 2b)**.

After filtering for disease mechanism, permissive evaluation of gene-phenotype fit and curation of pLoF variants, 131 variants remained **(Figure 2c).** Out of 131 variants, 60% (n = 78) were associated with neurodevelopmental conditions, 26% (n = 34) with neuromuscular conditions, 5% (n = 7) with ophthalmological conditions, 3% (n = 4) with renal conditions, and the remaining 5% with cardiovascular conditions (n = 3), musculoskeletal conditions (n = 2), craniofacial conditions (n = 1), hematologic conditions (n = 1), and reproductive conditions (n = 1) **(Figure 2d)**. This distribution is similar to that of the Broad CMG cohort overall, which is predominantly composed of neurodevelopmental and neuromuscular conditions with a smaller number of conditions involving other organ systems.

### Re-evaluation yielded 30 inherited diagnostic variants and six new diagnoses

The 131 inherited candidate variants were re-evaluated by 24 Broad CMG collaborating research teams to assess the diagnostic fit for the proband’s condition and to re-phenotype the parent. Of 131 variants, 62% (81 variants in 81 families) were determined by the respective domain expert to have sufficient gene-phenotype overlap to warrant further clinical assessment. For the remaining 38%, variants were considered a poor fit for the phenotype or further assessment of the variant-carrying parent and the proband was not possible **(Supplemental Figures 3a-b)**.

Of 81 variants, 30 variants in 30 families were considered diagnostic: the identified variant accounted for all or a portion of phenotypes of the enrolled proband **(Figure 3a-b)**. Assessment of the 30 transmitting parents in these families revealed that 50% (n = 15) showed IP with no discernible features and 20% (n = 6) were found to have mild features of the condition, supporting VE. Notably, following a deep clinical re-evaluation of parental phenotypes,13% (n = 4) were recognized as affected by the condition (complete penetrance). Additionally, for 17% (n = 5) of the probands, the inherited variant explained additional clinical features beyond those accounted for by another diagnostic variant (dual diagnosis). Four of these dual diagnosis cases had no signs of that phenotype in the parent (IP, dual diagnosis), whereas for 1 case the parent was later identified as affected (complete penetrance, dual diagnosis). When stratified by variant types, the final set consisted of 13 LoF, 12 missense, 4 Not LoF (protein truncating) but with a proximal P/LP ClinVar variant, and one synonymous variant predicted to impact splicing. Twenty (66%) had been previously reported as P/LP in ClinVar (**Supplemental Figure 4a).** IP was observed across all variant types (**Supplemental Figure 4b)**. Of the final 30 families with diagnoses **(Supplemental Table 2)**, six had been missed during the initial analysis. Among the six missed diagnoses, two were categorized as IP, two as VE, and two as complete penetrance. IP variants accounted for the diagnosis of 19 families of 4,325 families studied (0.4%). With the current solve rate of 26.9% in Broad CMG, a cohort where families have typically had prior genetic testing before CMG research sequencing, the 19 inherited incompletely penetrant variants account for 1.6% of all genetic diagnoses. Of note, genes for all 25 IP/VE families identified through this workflow had been previously linked to IP/VE in the literature **(Supplemental Table 2)**.

**Figure 3:**
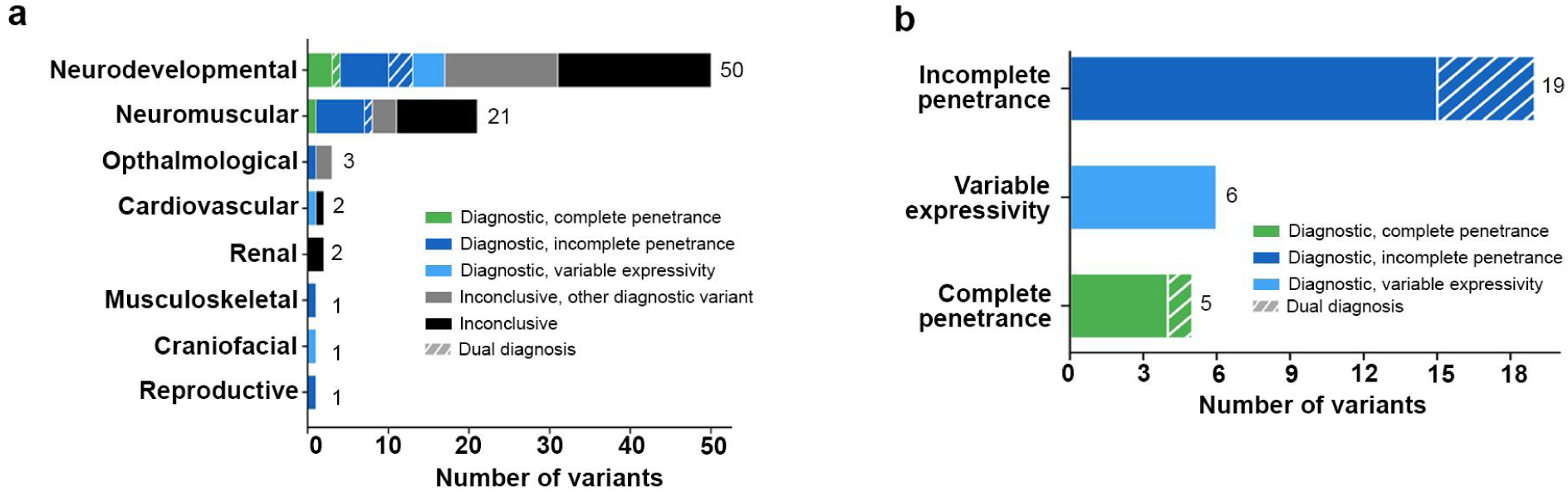
Phenotypic re-evaluation of 81 inherited putative disease-causing variants identified 30 variants as diagnostic. **(a)** Barplot depicting 81 putative disease-causing per disease group, colored by diagnostic outcome. **(b)** Barplot of 30 diagnostic inherited variants in 30 families showing the number of variants per penetrance category.

### Omics investigation of IP/VE due to an inherited *CHD8* frameshift variant

We performed omics follow-up of a family (RGP_526) in which a reportedly unaffected mother had transmitted an LoF frameshift variant (NM_001170629.2, chr14:21412981 T>TA, c.2157_2158insT and p.Asn720Ter) in *CHD8* to her affected offspring*. CHD8* haploinsufficiency causes an intellectual developmental disorder with autism and macrocephaly (OMIM #615032) and has a definitive gene-disease validity classification for complex neurodevelopmental disorder by ClinGen.^37^ The male proband (age of onset: 0-5 years) presented with macrocephaly, tall stature, high palate, autism, and global developmental delay, including delayed speech and language development since infancy (0-5 years) and is now in his early 20s. While detailed clinical re-evaluation of the mother confirmed that she was largely asymptomatic, she did exhibit subtle features, including minor learning difficulties during childhood and a head circumference at the upper end of the normal range. To remain stringent in our classification of IP, she was classified as VE **(Supplemental Table 2, Figure 4a, Supplemental Table 3)**.

**Figure 4:**
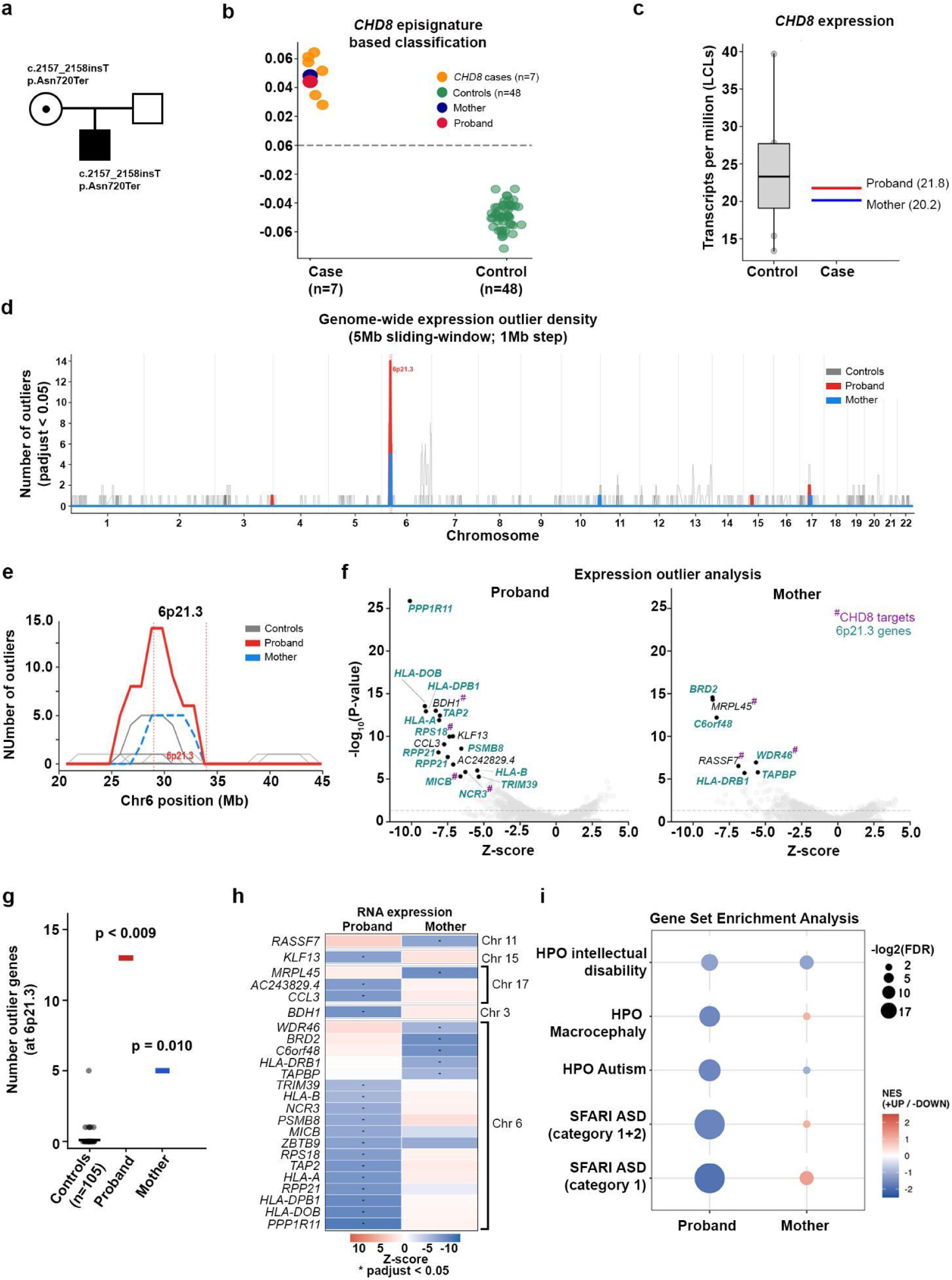
Omics characterization of IP/VE of an inherited *CHD8* frameshift variant. **(a)** Pedigree showing the proband inherited frameshift variant in *CHD8* gene from the largely asymptomatic mother. **(b)** *CHD8* episignature classification based on Pearson correlation. **(c)** Boxplot displaying *CHD8* expression in lymphoblast cell lines (LCLs) of proband, mother, and internal LCL controls (n = 5). **(d)** Expression outlier density using a 5 Mb sliding window with number of outliers in proband (red), mother (blue), compared with 105 controls (grey) genome-wide and **(e)** at the chromosome 6p21.3 locus. **(f)** Volcano plots showing LCL RNA expression outliers in the proband and the mother relative to the internal reference dataset. Genes on chr6p21.3 are marked in teal. The purple hash (#) denotes gene loci that are known *CHD8* targets. The dashed line indicates a transcriptome-wide significance threshold of 0.05. **(g)** Number of 6p21.3 gene expression outliers in controls, proband and mother ( p < 0.009 for proband compared to controls; p < 0.010 for mother compared to controls). **(h)** Heatmap displaying OUTRIDER Z-scores for gene expression outliers (p < 0.05) identified in proband and mother, sorted by chromosomal position. Blue blocks with asterisks (*) denotes significant downregulation, while orange/red blocks denote upregulation. **(i)** Dot plot displays normalized enrichment scores (NES) across high-confidence neurodevelopmental and ASD-associated gene sets derived from LCL RNA-seq. Evaluated categories encompass Human Phenotype Ontology (HPO) terms and curated SFARI autism risk categories. Circle size denotes statistical significance (−log_2_(FDR)) and color indicates directional enrichment (blue: downregulated; red: upregulated).

The proband’s diagnosis was further confirmed with a *CHD8* episignature analysis (svm = 0.81); notably, the mother also tested positive for the episignature (svm = 0.79).^42^ The findings were replicated using a previously described Pearson correlation-based classification score^43^ across *CHD8* episignature probes for proband, mother, *CHD8* cases (n = 7) and controls (n = 48) from previously published cases.^42^ Both proband and mother clustered closely with *CHD8* cases **(Figure 4b)**. A principal component analysis (PCA) revealed that while both the proband and mother’s data clustered on the *CHD8* case side of PC1, the mother mapped further away from the *CHD8* cases and the proband, suggesting distinct methylation patterns in the mother and the proband **(Supplemental Figure 5)**.

Transcriptomic analysis showed a similar reduction in *CHD8* expression in both the proband (transcripts per million [TPM] = 21.8) and the mother (TPM = 20.16) relative to the internal LCL reference data (median TPM = 27; **Figure 4c**). In the proband, 20% of RNA-seq reads contain the variant (26 of 130, p < 0.001, exact binomial test), compared to 18% of reads in the mother (23 of 130, p < 0.001), indicating allelic imbalance accounts for the reduction in *CHD8* expression, possibly due to NMD. However, the degree of allelic imbalance is not significantly different between the proband and mother (p = 0.75, Fisher’s exact test). Analysis of transcriptome-wide outlier expression in the proband and mother highlighted downregulation of the 6p21.3 locus in both individuals **(Figure 4d-e).** Downregulation of a gene cluster at chromosome 6p21.3 was seen in both the proband and the mother relative to the controls, but with more genes in the cluster showing significant transcriptome-wide expression in the proband compared to the mother (proband: 13 genes, p < 0.009 and mother: 5 genes, p = 0.010; **Figure 4f-g**). While both proband and mother had gene expression outliers on 6p21.3, each showed distinct outliers within this locus **(Figure 4h)**, several of these gene expression outliers are in known CHD8 genomic targets^49^: *BDHI, RPS18*, *MICB* and *NCR3* in proband, and *MRPL45*, *RASSF7* and *WDR46* in mother **(Figure 4h)**.

Gene set enrichment analysis (GSEA) of outlier expression genes demonstrated significant downregulation across autism and macrocephaly gene sets in proband, but not in the mother. The enriched gene sets in proband include high confidence SFARI Category 1 autism genes (n = 244, NES = −1.91, FDR = 4.5 x10^-6^), SFARI Category (1+2) autism gene set (n = 952, NES =−1.62, FDR = 4.4x10^-6^), HPO Autism (n= 207, NES =−1.59, FDR = 4.5x10^-3^) and HPO Macrocephaly (n= 112, NES =−1.58, FDR = 9.8 x10^-3^) networks **(Figure 4i)**. The HPO Intellectual Disability gene set demonstrated a similar trend but was not significant (n= 738, NES = −1.20, FDR = 0.072).

Lastly, to investigate the polygenic contribution to IP/VE, we calculated the polygenic risk score (PRS) for autism spectrum disorder (PGS000327)^48^ relative to ancestry matched individuals from the 1000 Genomes Project Phase 3. PRS for both mother and proband were observed to be below the population mean, which is consistent with previous reports demonstrating a reduction in PRS among families carrying high-impact monogenic risk.^24^ The unaffected mother scored lower (−1.23 SD) than the affected proband (−0.79 SD) **(Supplemental Figure 6)**.

## Discussion

*De novo* variants account for the majority of autosomal dominant rare disorders. However, we demonstrate that an evaluation for clinically relevant inherited variants transmitted from apparently asymptomatic or mildly symptomatic parents can increase diagnostic yield. Within our rare disease cohort, an evaluation for inherited variants identified 30 diagnostic variants in 30 families, 19 of which were of IP, 6 of which were of VE, and 5 were of complete penetrance. Of these 30 diagnostic variants, six resulted in new diagnoses that had been overlooked in previous analyses due to their inheritance, a lack of detailed phenotypes of transmitting parents, or insufficient disease evidence at the time of evaluation. Notably, cases of IP/VE had been previously reported for all 25 conditions identified in our cohort, though these conditions generally have high penetrance. The proportion of diagnostic variants inherited from a parent with IP in this cohort (1 in 60) may be slightly underestimated, as some of the 131 families were excluded from the re-evaluation due to limited opportunities for re-phenotyping. These findings underscore the importance of considering inherited IP variants in rare disease diagnostics, but also that this phenomenon is generally rare given the majority of these genes are associated with high penetrance.

To investigate the presence of differential omics signatures in an asymptomatic/mildly affected parent compared with the affected proband, we performed epigenomic and transcriptomic analyses of the *CHD8* family. The proband, as well as the reportedly asymptomatic/mildly affected mother, tested positive for the *CHD8*-disease episignature. Principal component analysis identified a divergent epigenomic profile in the mother compared with the proband and other *CHD8* cases, indicating the IP/VE could potentially be reflected in the methylation profile. A transcriptomic study of LCLs revealed a similar degree of reduced *CHD8* expression in both proband and mother, suggesting that compensatory wild-type expression in the mother is not the cause of IP/VE. Transcriptome-wide outlier analysis showed overlap with genes harboring *CHD8* binding sites^49^, with a distinct set of target genes downregulated in proband versus mother. Expression outlier genes clustered to a specific region, chromosome 6p21.3, in both individuals. Interestingly, the mother exhibited a less extensive dysregulation of 6p21.3 (5 genes versus 13 in proband). The 13 6p21 genes with reduced expression identified in the proband included genes reported to be associated with neurodevelopmental risk, such as *PPP1R11,*^52,53^ and *HLA-A*, *HLA-DPB1* and *HLA-DOB* genes implicated in ASD susceptibility and immune dysfunction.^54–56^ While this genomic region has a plausible biological relevance to ASD, further functional validation is required to establish this mechanism conclusively. GSEA implicated downregulation across autism, macrocephaly, and neurodevelopmental gene sets in the proband but not in the mother, pointing to a shift that could contribute to the phenotypic variability. Lastly, we tested the burden of common ASD risk variation using established PRS. A nominally lower PRS was observed in the mother, reflecting a higher burden of potentially protective common alleles that could modify penetrance. However, given the modest difference observed here, the result should be interpreted cautiously^24^.

In summary, our findings demonstrate that systematic evaluation of parentally inherited variants can identify clinically relevant diagnoses that would otherwise be missed and should be considered as part of rare disease diagnostic workflows. Integrating genomic, transcriptomic, and epigenomic data further provides a framework for investigating the molecular basis of incomplete penetrance and variable expressivity, ultimately improving variant interpretation and diagnostic precision.

## Data Availability

Genomic and phenotypic data from the Broad CMG is available via dbGaP accession numbers phs003047 (GREGoR) and phs001272 (CMG).

## Supporting information

Revised_Supplemental Tables-Sytematic_evaluation_of _incomplete_penetrance_in_4325_rare_disease_CMG_families_singh etal

## Acknowledgments

We thank all the families who participated in the study. We are thankful to all the Broad CMG collaborating laboratories and investigators, including Dr. Sander Pajusalu, Dr. Katrin Õunap, Dr. Karit Reinson, Dr. Gina O’ Grady, Dr. Laura St Clair, Dr. Alan Ma, Dr. Wendy Chung, Dr. Friedhelm Hildebrandt, Dr. Shirlee Shril, Dr. Sandra Cooper, Dr. Rhett Merchant, Dr. Leigh Waddell, Dr. Eric Pierce, Dr. Kinga Bujakowska and Dr. Emily Place, for providing valuable details and multiple reviews for their respective families. This publication is based on research supported by The G. Harold and Leila Y. Mathers Charitable Foundation. The Genotype-Tissue Expression (GTEx) Project was supported by the Common Fund of the Office of the Director of the National Institutes of Health, and by NCI, NHGRI, NHLBI, NIDA, NIMH, and NINDS.

## Author Contributions

Conceptualization: S.G. and A.O.D.L.; Curation: M.S., S.S., M.S.B., E.G. and S.G; Formal Analysis: M.S., S.S, J.M., S.C., E.O.H., S.D.T., I.O.O, L.P., G.L., M.O.L., C.A.T, C.A.G., K.N.A., M.W., A.H.B., J.E.N., A.L., C.A.W., J.A.J., B.J.B, E.C.E., S.D., C.G.B., E.A., E.H.S., P.K., Z.B., K.A.D., V.S., A.T, S.M.W., T.Y.T., L.G., P.A., M.R.J., T.T.H, C.P.B., A.B.B., H.S.S., V.S.G., H.L.R., R.W. A.O.D.L and S.G.; Funding Acquisition: S.G. and A.O.D.L; Investigation: M.S.; Visualization: M.S., Supervision: S.G. and A.O.D.L. Writing-original draft: M.S. and S.G Writing-review & editing: S.G. A.O.D.L., H.L.R, S.S., V.S.G., J.A.J., E.C.E, A.H.B., M.W., J.E.N. and M.R.J. All authors reviewed and approved the final draft of the manuscript.

## Ethics Declaration

All participants provided informed consent to research participation. This study was approved by the Mass General Brigham Institutional Review Board (protocols 2016P001422 and 2013P001477).

## Supplementary Figures

**Supplemental Figure 1:**
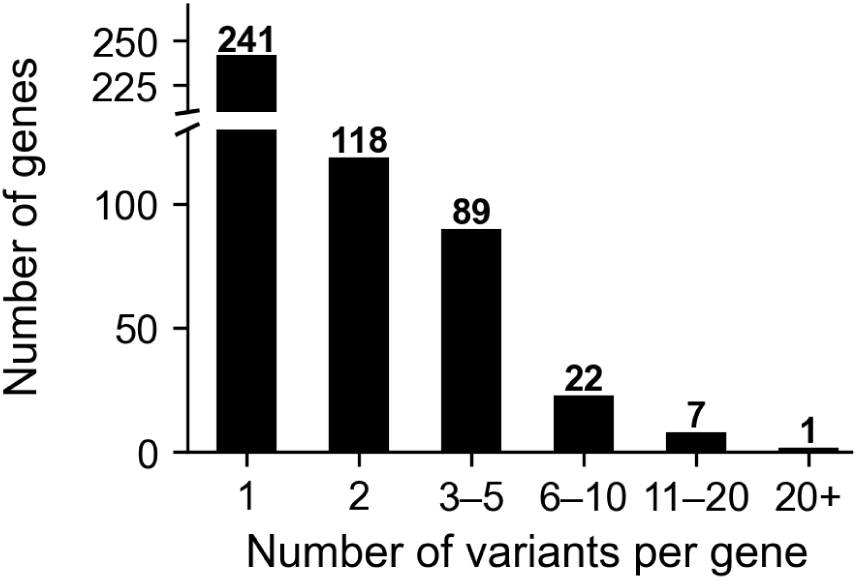
Variant counts across genes. Plot showing number of variants per gene. Of 478 genes, 50.4% (n = 241) harbored a single variant, 24.7% (n = 118) harbored two variants, 18.6% (n = 89) harbored 3-5 variants, 4.6% (n = 22) harbored 6-10 variants, 1.5% (n = 7) harbored 11-20 variants, and 0.2% (n = 1) harbored more than 20 variants.

**Supplemental Figure 2:**
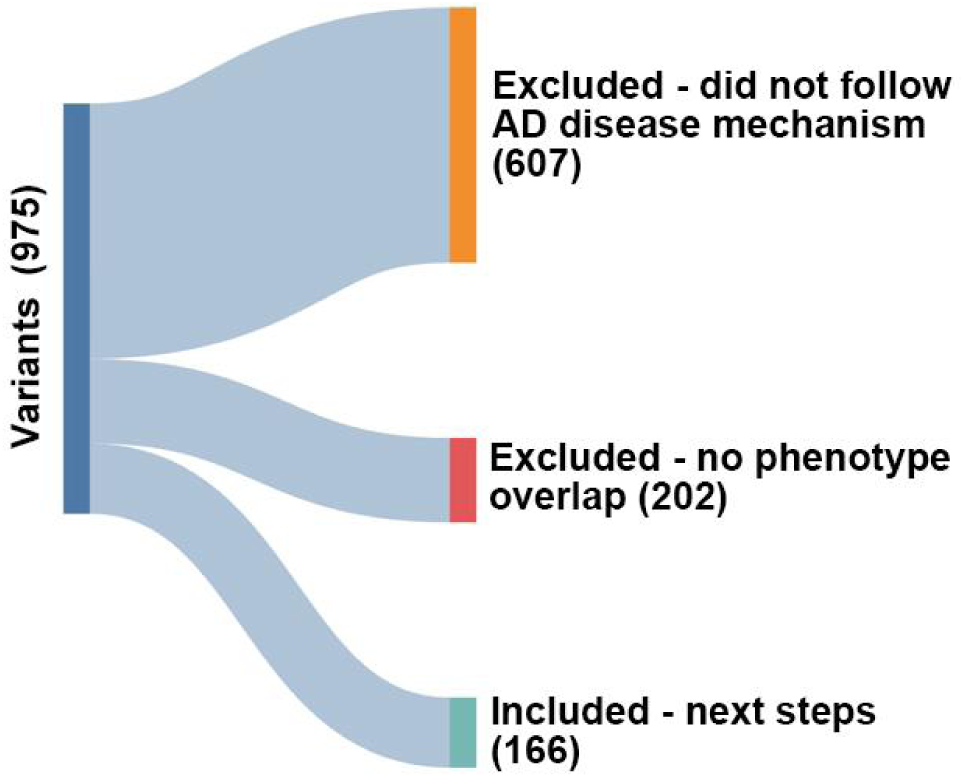
Sankey plot showing distribution of excluded and included variants after filtering for disease mechanism and gene-phenotype overlap. Sankey plot shows 62% (n = 607) of the variants from initial set were excluded because they did not follow autosomal dominant (AD) mechanism of disease, 21% (n = 202) were excluded due to lack of gene-phenotype overlap and 17% (n = 166) of variants were included for further analysis.

**Supplemental Figure 3:**
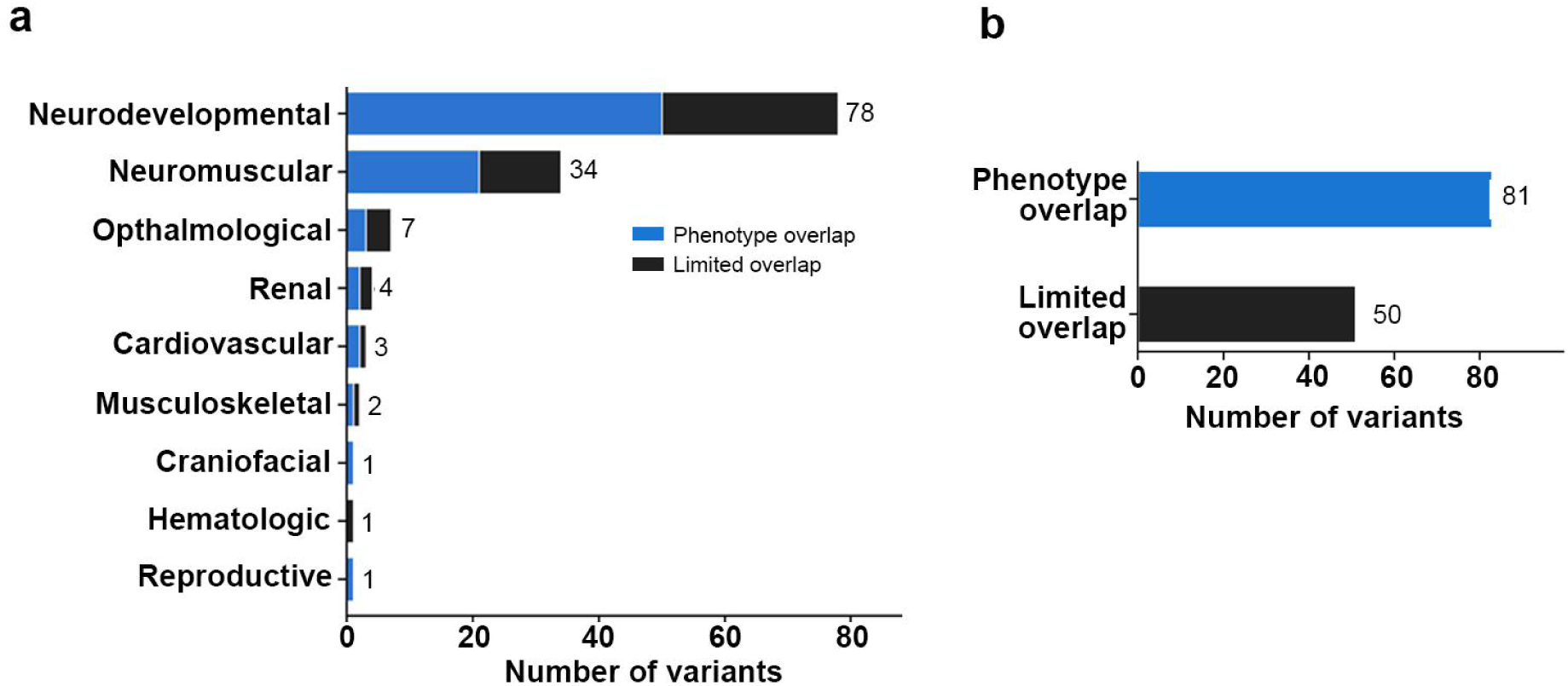
Re-phenotyping evaluation of variants for diagnostic fit of 131 candidate variants. **(a)** Barplot of 131 candidate variants that were putative disease-causing across disease groups. **(b)** Phenotypic overlap was found in 62% (n = 81) and were classified as putative disease-causing.

**Supplemental Figure 4:**
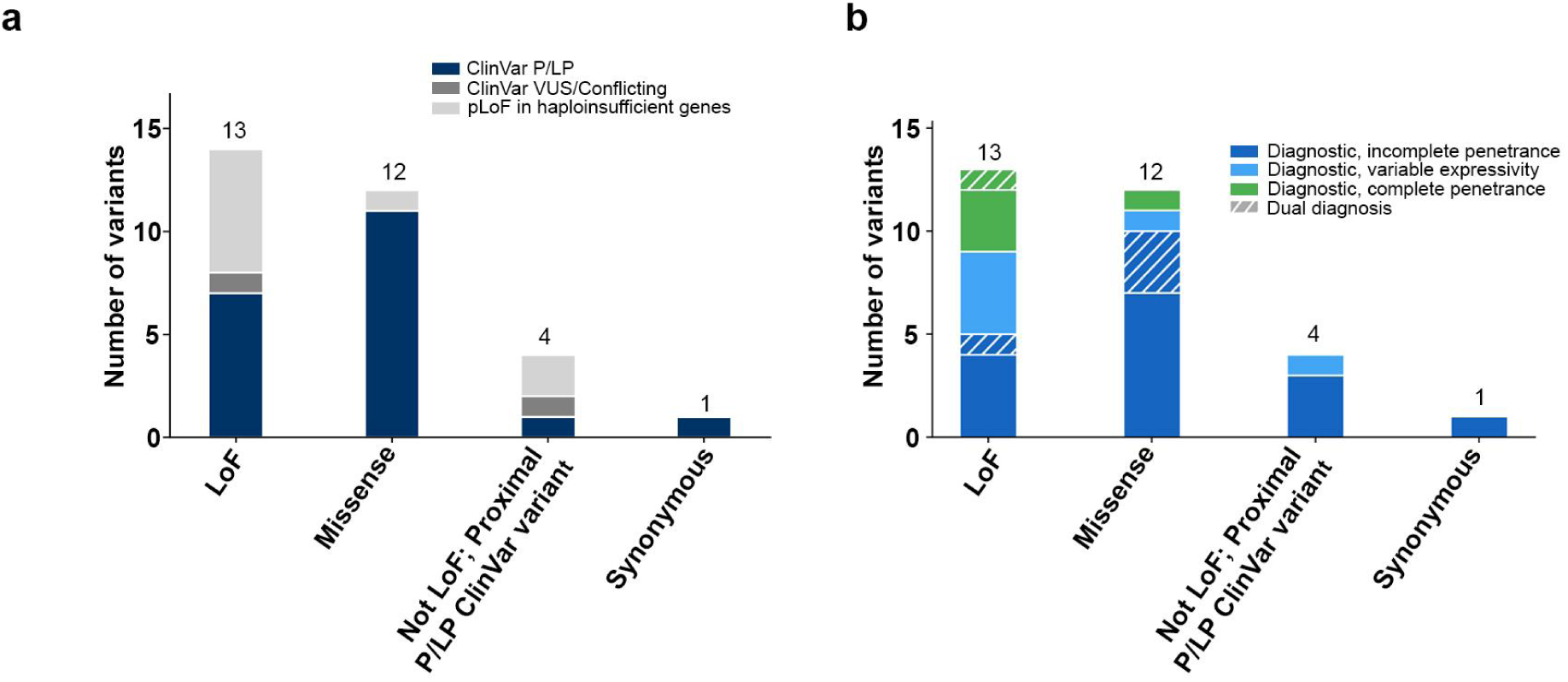
Variant types in the 30 diagnostic families. **(a)** Barplot of 30 diagnostic variants per variant type, colored by P/LP: pathogenic/likely pathogenic; VUS: variant of uncertain significance; pLoF: predicted loss of function variant. **(b)** Barplot of 30 diagnostic variants per variant type, colored by penetrance category.

**Supplemental Figure 5:**
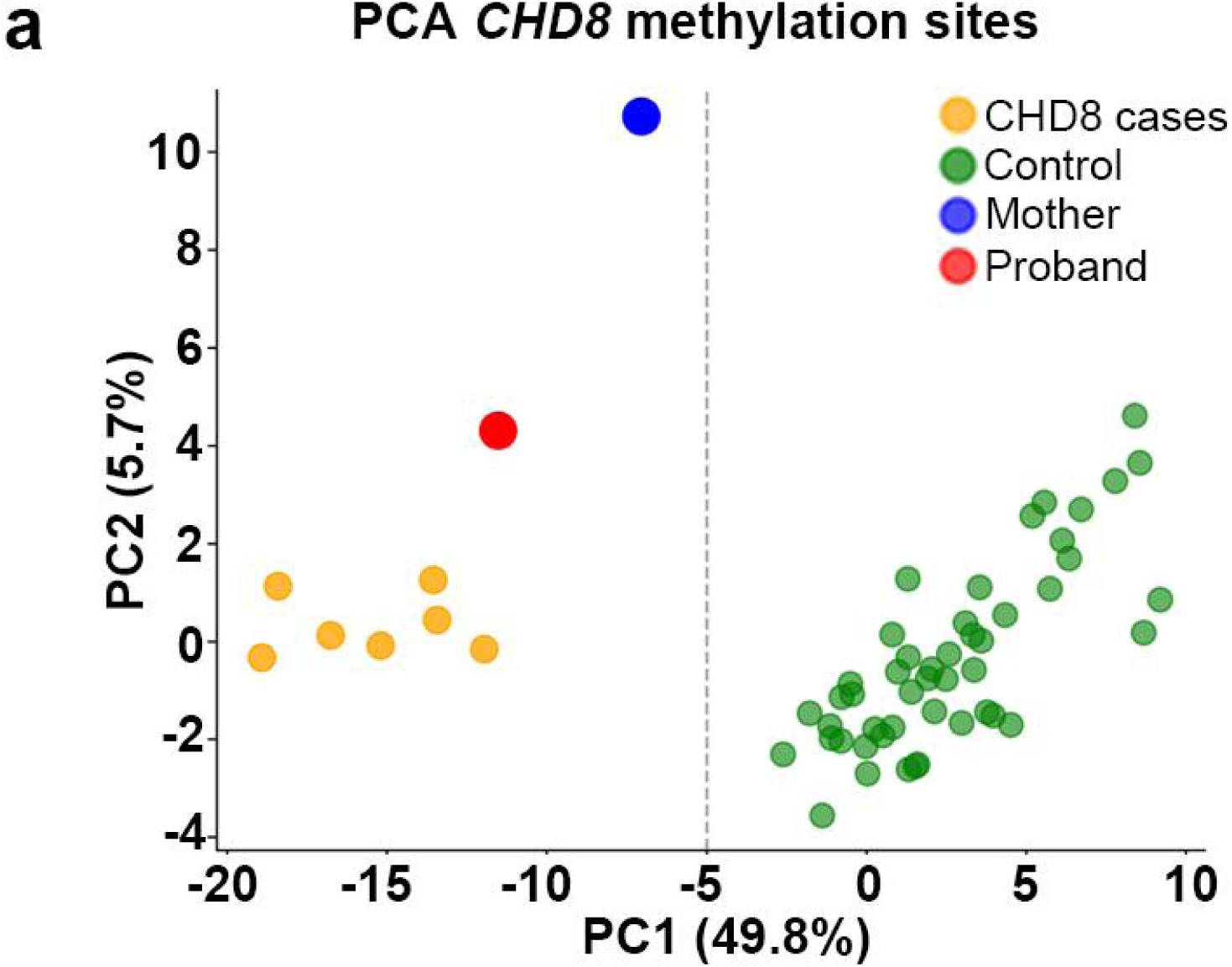
PCA of DNA methylation beta values. The dashed grey line separates cases from controls. PC1 explains maximum (49.8%) of the variance in the data and separates controls from cases and the *CHD8* family (mother and proband).

**Supplemental Figure 6:**
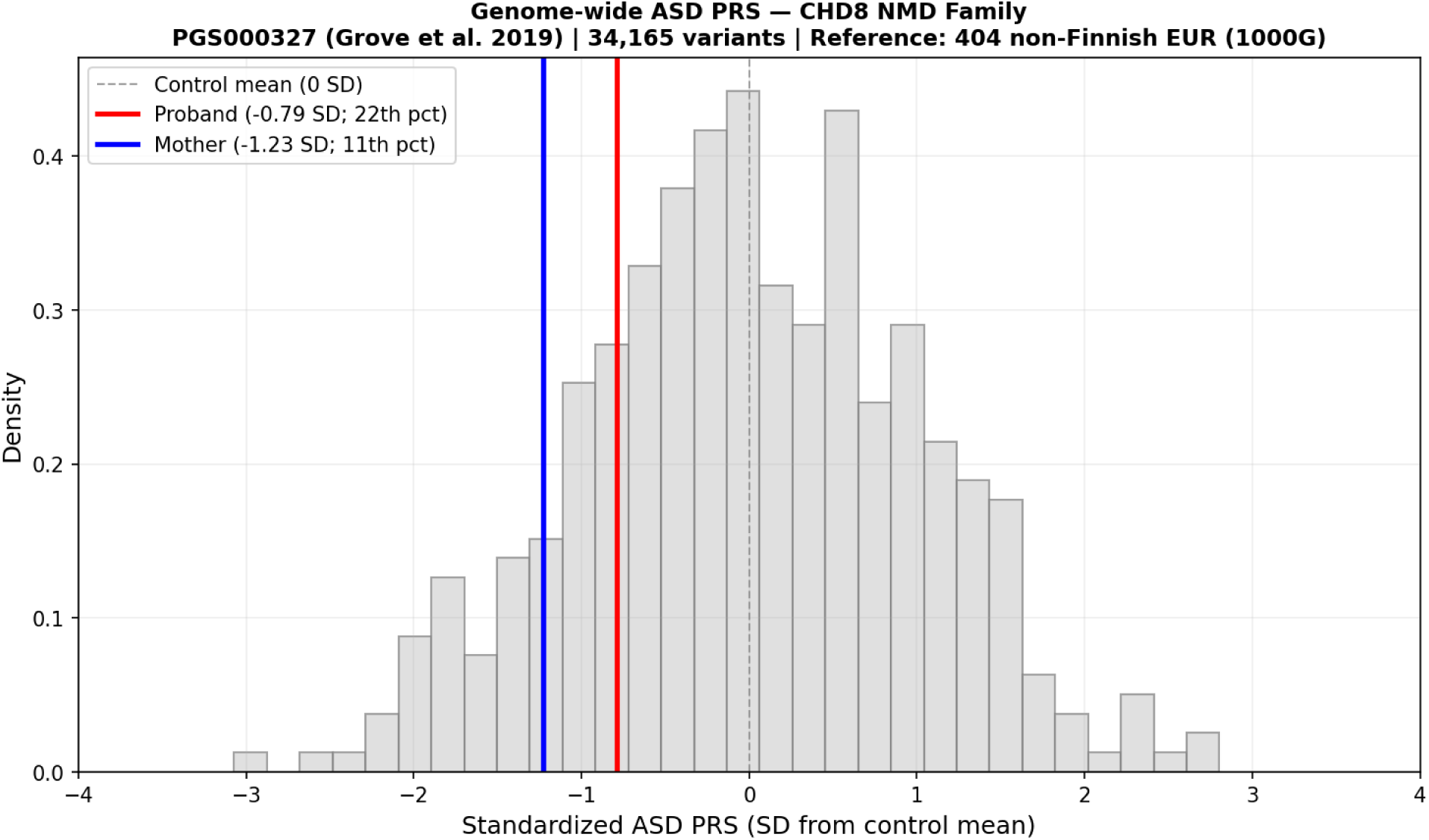
Assessment of Autism Spectrum Disorder (ASD) variant burden using ASD polygenic risk scores (PRS) in the *CHD8* family. Histogram plot of standardized ASD PRS in ancestry matched individuals sourced from 1000 Genomes Project Phase 3 (grey bars). Control scores are normalized to a control mean of 0 (dashed line) and SD = 1. Proband 22th percentile (red) and mother 11th percentile (blue) with SD values and percentile noted in the legend top left box). Genome-wide ASD PRS calculated using 35,087 variants from PGS000327 (Grove et al., 2019).

## References

1. Wright CF, Campbell P, Eberhardt RY, et al. Genomic diagnosis of rare pediatric disease in the United Kingdom and Ireland. N Engl J Med. 2023;388(17):1559–1571.

2. Chen Y, Dawes R, Kim HC, et al. De novo variants in the non-coding spliceosomal snRNA gene RNU4-2 are a frequent cause of syndromic neurodevelopmental disorders. medRxiv. Published online April 9, 2024:2024.04.07.24305438.

3. Wojcik MH, Lemire G, Berger E, et al. Genome Sequencing for Diagnosing Rare Diseases. N Engl J Med. 2024;390(21):1985–1997.

4. Stenton SL, O’Leary M, Lemire G, et al. Critical assessment of variant prioritization methods for rare disease diagnosis within the Rare Genomes Project. medRxiv. Published online August 4, 2023. doi:10.1101/2023.08.02.23293212

5. Rehm HL. Evolving health care through personal genomics. Nat Rev Genet. 2017;18(4):259–267.

6. Clark MM, Stark Z, Farnaes L, et al. Meta-analysis of the diagnostic and clinical utility of genome and exome sequencing and chromosomal microarray in children with suspected genetic diseases. NPJ Genom Med. 2018;3:16.

7. Gelb BD. 2024 ASHG presidential address: Incomplete penetrance and variable expressivity: Old concepts, new urgency. Am J Hum Genet. 2025;112(3):461–466.

8. Kingdom R, Wright CF. Incomplete Penetrance and Variable Expressivity: From Clinical Studies to Population Cohorts. Front Genet. 2022;13:920390.

9. Minikel EV, Vallabh SM, Lek M, et al. Quantifying prion disease penetrance using large population control cohorts. Sci Transl Med. 2016;8(322):322ra9.

10. Tuke MA, Ruth KS, Wood AR, et al. Mosaic Turner syndrome shows reduced penetrance in an adult population study. Genet Med. 2019;21(4):877–886.

11. Sharp LN, Colclough K, Murray Leech J, et al. Population Prevalence, Penetrance, and Mortality for Genetically Confirmed MODY. J Clin Endocrinol Metab. 2026;111(5):e1388–e1395.

12. Goodrich JK, Singer-Berk M, Son R, et al. Determinants of penetrance and variable expressivity in monogenic metabolic conditions across 77,184 exomes. Nat Commun. 2021;12(1):3505.

13. Zhytnik L, Maasalu K, Pashenko A, et al. Pathogenic Variants and Phenotype Characteristics in Ukrainian Osteogenesis Imperfecta Patients. Front Genet. 2019;10:722.

14. Pagnamenta AT, Fasham J, Beaumont RN, et al. Reduced penetrance of *COL1A1/2* pathogenic variants linked with osteogenesis imperfecta: analysis of a large population cohort. medRxiv. Published online October 23, 2025:2025.10.22.25338523. doi:10.1101/2025.10.22.25338523

15. Alkaf B, Jha A, Saad A, et al. The Emirati Genome Program enables population-wide penetrance estimation and novel discovery for inherited retinal disease. medRxiv. Published online October 19, 2025:2025.10.14.25337784. doi:10.1101/2025.10.14.25337784

16. Zaslavsky K, Chen L, Park C, et al. Low population penetrance of variants associated with inherited retinal degenerations. Am J Hum Genet. 2026;113(1):71–82.

17. Forrest IS, Chaudhary K, Vy HMT, et al. Population-Based Penetrance of Deleterious Clinical Variants. JAMA. 2022;327(4):350–359.

18. Wright CF, West B, Tuke M, et al. Assessing the Pathogenicity, Penetrance, and Expressivity of Putative Disease-Causing Variants in a Population Setting. Am J Hum Genet. 2019;104(2):275–286.

19. Gudmundsson S, Singer-Berk M, Stenton SL, et al. Exploring penetrance of clinically relevant variants in over 800,000 humans from the Genome Aggregation Database. Nature Communications. 2025;16(1):9623.

20. Tarailo-Graovac M, Zhu JYA, Matthews A, van Karnebeek CDM, Wasserman WW. Assessment of the ExAC data set for the presence of individuals with pathogenic genotypes implicated in severe Mendelian pediatric disorders. Genet Med. 2017;19(12):1300–1308.

21. Khan A, Shang N, Nestor JG, et al. Polygenic risk affects the penetrance of monogenic kidney disease. medRxiv. Published online May 10, 2023. doi:10.1101/2023.05.07.23289614

22. Downs B, Sherman S, Cui J, et al. Common genetic variants contribute to incomplete penetrance: evidence from cancer-free BRCA1 mutation carriers. Eur J Cancer. 2019;107:68–78.

23. Alper CA, Awdeh Z. Incomplete penetrance of MHC susceptibility genes: prospective analysis of polygenic MHC-determined traits. Tissue Antigens. 2000;56(3):199–206.

24. Huang QQ, Wigdor EM, Malawsky DS, et al. Examining the role of common variants in rare neurodevelopmental conditions. Nature. 2024;636(8042):404–411.

25. Molinari E, Nickerson A, Quistrebert J, Kless A. High Frequency of Repeat Expansions in UK Biobank Suggests Protective Rare Variants Modulate C9ORF72 Penetrance in Neurodegeneration. medRxiv. Published online September 19, 2025:2025.09.18.25336048. doi:10.1101/2025.09.18.25336048

26. Xu J, Zou Z, Liu W, et al. HAPLN3 p.T34A contributes to incomplete penetrance of moyamoya disease in Chinese carrying RNF213 p.R4810K. Eur J Neurol. 2024;31(12):e16473.

27. Kingdom R, Beaumont RN, Wood AR, Weedon MN, Wright CF. Genetic modifiers of rare variants in monogenic developmental disorder loci. Nat Genet. 2024;56(5):861–868.

28. Corty RW, Brogan J, Byram K, Springer J, Grayson PC, Bick AG. VEXAS-Defining UBA1 Somatic Variants in 245,368 Diverse Individuals in the NIH All Of Us Cohort. Arthritis Rheumatol. 2024;76(6):942–948.

29. Stewart O’jay, Gruber C, Randolph HE, et al. Monoallelic expression can govern penetrance of inborn errors of immunity. Nature. 2025;637(8048):1186–1197.

30. Li D, March ME, Wang T, et al. Exome and RNA-Seq analyses of an incomplete penetrance variant in USP9X in female-specific syndromic intellectual disability. Am J Med Genet A. 2022;188(6):1808–1814.

31. Einson J, Glinos D, Boerwinkle E, et al. Genetic control of mRNA splicing as a potential mechanism for incomplete penetrance of rare coding variants. bioRxiv. Published online January 31, 2023. doi:10.1101/2023.01.31.526505

32. Chen P, Zampawala Z, Wang H, Wang L. Exploring the impact of a KCNH2 missense variant on Long QT syndrome: insights into a novel gender-selective, incomplete penetrance inheritance mode. Front Genet. 2024;15:1409459.

33. Magri S, Nanetti L, Gellera C, et al. Digenic inheritance of STUB1 variants and TBP polyglutamine expansions explains the incomplete penetrance of SCA17 and SCA48. Genet Med. 2022;24(1):29–40.

34. Mao K, Borel C, Ansar M, et al. FOXI3 pathogenic variants cause one form of craniofacial microsomia. Nat Commun. 2023;14(1):2026.

35. Michaud V, Lasseaux E, Green DJ, et al. The contribution of common regulatory and protein-coding TYR variants to the genetic architecture of albinism. Nat Commun. 2022;13(1):3939.

36. Timberlake AT, Choi J, Zaidi S, et al. Two locus inheritance of non-syndromic midline craniosynostosis via rare and common alleles. Elife. 2016;5. doi:10.7554/eLife.20125

37. Rehm HL, Berg JS, Brooks LD, et al. ClinGen--the Clinical Genome Resource. N Engl J Med. 2015;372(23):2235–2242.

38. Pais LS, Snow H, Weisburd B, et al. seqr: A web-based analysis and collaboration tool for rare disease genomics. Hum Mutat. 2022;43(6):698–707.

39. Amberger JS, Bocchini CA, Schiettecatte F, Scott AF, Hamosh A. OMIM.org: Online Mendelian Inheritance in Man (OMIM®), an online catalog of human genes and genetic disorders. Nucleic Acids Res. 2015;43(Database issue):D789–D798.

40. Singer-Berk M, Gudmundsson S, Baxter S, et al. Advanced variant classification framework reduces the false positive rate of predicted loss-of-function variants in population sequencing data. Am J Hum Genet. 2023;110(9):1496–1508.

41. Jaganathan K, Kyriazopoulou Panagiotopoulou S, McRae JF, et al. Predicting Splicing from Primary Sequence with Deep Learning. Cell. 2019;176(3):535–548.e24.

42. Siu MT, Butcher DT, Turinsky AL, et al. Functional DNA methylation signatures for autism spectrum disorder genomic risk loci: 16p11.2 deletions and CHD8 variants. Clin Epigenetics. 2019;11(1):103.

43. Choufani S, Cytrynbaum C, Chung BHY, et al. NSD1 mutations generate a genome-wide DNA methylation signature. Nat Commun. 2015;6:10207.

44. Pedregosa F, Varoquaux G, Gramfort A, et al. Scikit-learn: Machine learning in Python. the Journal of machine Learning research. 2011;12:2825–2830.

45. GTEx Consortium. The GTEx Consortium atlas of genetic regulatory effects across human tissues. Science. 2020;369(6509):1318–1330.

46. Brechtmann F, Mertes C, Matusevičiūtė A, et al. OUTRIDER: A Statistical Method for Detecting Aberrantly Expressed Genes in RNA Sequencing Data. Am J Hum Genet. 2018;103(6):907–917.

47. Simons Foundation Autism Research Initiative. Gene Scoring Module - SFARI Gene. Accessed June 12, 2026. https://gene.sfari.org/database/gene-scoring/

48. Grove J, Ripke S, Als TD, et al. Identification of common genetic risk variants for autism spectrum disorder. Nat Genet. 2019;51(3):431–444.

49. Sugathan A, Biagioli M, Golzio C, et al. CHD8 regulates neurodevelopmental pathways associated with autism spectrum disorder in neural progenitors. Proceedings of the National Academy of Sciences. 2014;111(42):E4468–E4477.

50. Rockowitz S, Shao W, French C, et al. Scaling genomic reanalysis to unlock diagnoses and transform rare disease care. HGG Adv. 2026;7(2):100582.

51. Welland MJ, Ahlquist KD, De Fazio P, et al. Scalable automated reanalysis of genomic data in research and clinical rare disease cohorts. medRxiv. Published online May 21, 2025. doi:10.1101/2025.05.19.25327921

52. Cross-Disorder Group of the Psychiatric Genomics Consortium, Lee SH, Ripke S, et al. Genetic relationship between five psychiatric disorders estimated from genome-wide SNPs. Nat Genet. 2013;45(9):984–994.

53. Lotan A, Fenckova M, Bralten J, et al. Neuroinformatic analyses of common and distinct genetic components associated with major neuropsychiatric disorders. Front Neurosci. 2014;8:331.

54. Torres AR, Sweeten TL, Cutler A, et al. The association and linkage of the HLA-A2 class I allele with autism. Hum Immunol. 2006;67(4-5):346–351.

55. Lombardi L, Le Clerc S, Wu CL, et al. A human leukocyte antigen imputation study uncovers possible genetic interplay between gut inflammatory processes and autism spectrum disorders. Transl Psychiatry. 2023;13(1):244.

56. Guerini FR, Bolognesi E, Mensi MM, et al. HLA-A, -B, -C and -DRB1 Association with Autism Spectrum Disorder Risk: A Sex-Related Analysis in Italian ASD Children and Their Siblings. Int J Mol Sci. 2024;25(18). doi:10.3390/ijms25189879

